# Genome-Wide Determinants of Cellular Immune Responses to Mumps Vaccine

**DOI:** 10.1101/2023.04.27.23289213

**Authors:** Inna G. Ovsyannikova, Iana H. Haralambieva, Daniel J. Schaid, Nathaniel D. Warner, Gregory A. Poland, Richard B. Kennedy

## Abstract

**Background:** We have previously described genetic polymorphisms in candidate genes that are associated with inter-individual variations in antibody responses to mumps vaccination. To expand upon our previous work, we performed a genome-wide association study (GWAS) to discover host genetic variants associated with mumps vaccine-induced cellular immune responses.

**Methods:** We performed a GWAS of mumps-specific immune response outcomes (11 secreted cytokines/chemokines) in a cohort of 1,406 subjects.

**Results:** Among the 11 cytokine/chemokines we studied, four (IFN-γ, IL-2, IL-1β, and TNFα) demonstrated GWAS signals reaching genome-wide significance (p<5 x 10^-8^). A genomic region (encoding Sialic acid-binding immunoglobulin-type lectins/SIGLEC) located on chromosome 19q13 (p<5×10^-8^) was associated with both IL-1β and TNFα responses. The SIGLEC5/SIGLEC14 region contained 11 statistically significant single nucleotide polymorphisms (SNPs), including the intronic SIGLEC5 rs872629 (p=1.3E-11) and rs1106476 (p=1.32E-11) whose alternate alleles were significantly associated with decreased levels of mumps-specific IL-1β (rs872629, p=1.77E-09; rs1106476, p=1.78E-09) and TNFα (rs872629, p=1.3E-11; rs1106476, p=1.32E-11) production.

**Conclusions:** Our results suggest that SNPs in the SIGLEC5/SIGLEC14 genes play a role in cellular and inflammatory immune responses to mumps vaccination. These findings motivate further research into the functional roles of SIGLEC genes in the regulation of mumps vaccine-induced immunity.

## 1. Introduction

The widespread use of mumps vaccines, especially the two-dose immunization schedule introduced in 1989, has dramatically decreased but not eliminated the occurrence of mumps in the US. Despite widespread use of mumps vaccine, a resurgence of mumps occurred in the US from 2005 to date, with multiple outbreaks among highly immunized populations [1-3]. This is evidenced by the recent mumps resurgence and the largest outbreaks in decades in the 45 US states, which resulted in more than 10,000 cases [4, 5]. A recent CDC study found that during the 2007 to 2019 period, a total of 9,172 pediatric mumps cases were reported. Between 81% and 94% of these pediatric cases occurred among children who had received one or more doses of the mumps-containing vaccine [6]. The same CDC study found that one-third (range 13%-59%) of reported mumps cases in the US between 2007 and 2019 occurred among children and adolescents, most of whom were vaccinated [6].

Vaccine failure is a significant factor in these outbreaks, with most cases having received two previous doses of mumps vaccine [7, 8]. Mumps is not just a disease of children. For example, among French military recruits vaccinated five years previously with two doses of measles-mumps-rubella (MMR) vaccine, only 76% developed protective antibody titers against mumps compared to 96% for measles and 98% for rubella [9]. Among healthy contacts of cases on US college campuses, the effectiveness of one dose of mumps vaccine was 64% and for two doses only 76% [10]. In a 2005 study of 312 cases in England, effectiveness among older students 11-12 years of age was 66% for one dose and 86% for two doses [11]. Studies report vaccine effectiveness of 49% to 82% for one dose, and 66% to 88% for two doses of mumps vaccine [2, 5, 12, 13].

The reasons for this apparent vaccine failure are unknown, but critical to understand. Mumps immunity is influenced by several factors, including host genetics; however, our understanding of the role of these factors is limited. Mumps vaccine immunogenicity has been reliably assessed in multiple studies; however, immune response measures have been mostly confined to measuring antibody titers as an imperfect correlate of immunity [14]. Identifying genetic determinants of the adaptive immune response to mumps will inform our understanding of mumps immunity. To date there have been no population-based studies identifying associations between mumps vaccine-induced immune responses and genome-wide SNPs, despite the public health implications. We previously published a twins study demonstrating that the heritability (the ratio of genetic variance to total variance) of mumps vaccine Ab response was ∼40% [15]. Despite the relatively small cohort size (n=100) in that study, our results supported a genetic component to mumps vaccine response. We also previously described genetic polymorphisms in candidate genes that are associated with inter-individual variations in Ab responses to mumps vaccine [16]. These vaccine studies have primarily concentrated on the HLA region and were based on a small number of candidate genes/SNPs.

To expand upon our previous work, we performed a genome-wide association study (GWAS) to discover host genetic variants associated with inter-individual variation in mumps vaccine-induced cellular immune response outcomes. We report the first GWAS study of mumps vaccine-induced cellular (cytokine/chemokine) immune responses in a cohort of 1,406 subjects who received live attenuated mumps vaccine.

## 2. Methods

The immune assays and genotyping procedure described herein are similar or identical to those published for our previous vaccine studies [17-19].

### 2.1. Subjects

We analyzed a large sample of 1,406 healthy children, older adolescents, and healthy adults consisting of two combined Rochester and San Diego cohorts (Table 1). The Rochester cohort consisted of healthy subjects (n=748; 11–19 years of age) who received two doses of the mumps-containing vaccine as the live viral vaccine licensed in the US. The San Diego cohort consisted of healthy subjects (n=658; 19–40 years of age) from the US Armed Forces who either demonstrated proof of past mumps vaccination at the time of enlistment in the military service or received MMR vaccination at that time. The demographic and clinical characteristics of these cohorts have been previously published [17, 18, 20, 21]. Due to constraints on PBMC sample availability and the small number of minority groups in the cohort, this study focused on individuals with European/Asian ancestry.

**Table 1.** Demographic characterization of the study subjects.

| **Table 1.** Demographic characterization of the study subjects. |  |  |  |
| --- | --- | --- | --- |
|  | **Rochester cohort**<br>**(n=748)** | **San Diego cohort**<br>**(n=658)** | **Total (n=1,406)** |
| **Age at enrollment**<br>**(years)** |  |  |  |
| Mean (SD) | 14.918 (2.215) | 24.767 (4.235) | 19.528 (5.930) |
| Range | 11.000 - 19.000 | 19.000 - 40.000 | 11.000 - 40.000 |
| Interquartile range (IQR) | 13.0 - 17.0 | 22.0 – 27.0 | 15.0 – 23.0 |
| **Sex** |  |  |  |
| Female | 327 (43.7%) | 176 (26.7%) | 503 (35.8%) |
| Male | 421 (56.3%) | 482 (73.3%) | 903 (64.2%) |
| **Self-reported ancestry** |  |  |  |
| American Indian/Alaska native | 4 (0.5%) | 16 (2.4%) | 20 (1.4%) |
| Asian/Hawaiian/Pacific Islander | 22 (2.9%) | 39 (5.9%) | 61 (4.3%) |
| Multiple | 23 (3.1%) | 68 (10.3%) | 91 (6.5%) |
| Other | 4 (0.5%) | 106 (16.1%) | 110 (7.8%) |
| Unknown | 5 (0.7%) | 19 (2.9%) | 24 (1.7%) |
| White | 690 (92.2%) | 410 (62.3%) | 1100 (78.2%) |
| **STRUCTURE-determined ancestry** |  |  |  |
| Asian | 20 (2.7%) | 59 (9.0%) | 79 (5.6%) |
| EUR | 728 (97.3%) | 599 (91.0%) | 1327 (94.4%) |
| **Self-reported ethnicity** |  |  |  |
| Don't Know | 5 (0.7%) | 17 (2.6%) | 22 (1.6%) |
| Hispanic | 15 (2.0%) | 181 (27.5%) | 196 (13.9%) |
| Not Hispanic | 728 (97.3%) | 460 (69.9%) | 1188 (84.5%) |
| **Age at first MMR**<br>**(months)** |  |  |  |
| Number missing | 0 | 658 | 658 |
| Mean (SD) | 16.689 (9.704) | NA | 16.689 (9.704) |
| Range | 11.000 - 185.000 | NA | 11.000 - 185.000 |
| **Age at last MMR (years)** |  |  |  |
| Number missing | 0 | 184 | 184 |
| Mean (SD) | 8.390 (3.494) | 20.359 (3.169) | 13.033 (6.738) |
| Range | 1.000 - 17.000 | 17.000 - 35.000 | 1.000 - 35.000 |
| **Time from last MMR to blood draw (years)** |  |  |  |
| Number missing | 0 | 184 | 184 |
| Mean (SD) | 6.526 (2.762) | 3.310 (1.607) | 5.279 (2.851) |
| Range | 0.400 - 15.500 | 0.300 - 8.600 | 0.300 - 15.500 |

The Institutional Review Boards of the Mayo Clinic (Rochester, MN) and the NHRC (Naval Health Research Center, San Diego, CA) approved the study, and written informed consent was obtained from each subject.

### 2.2. Cytokines and chemokines secretion

We measured the cytokine (IL-2, IL-6, IL-10, IFNα2a, IFNγ, IL-1β, and TNFα) and chemokine (IP-10, MCP-1, MIP-1α, and MIP-1β) response in PBMCs (cell culture supernatants were tested in duplicate) from 1,406 subjects with/without stimulation with mumps virus antigen (Enders strain, Bio-Rad/Abd Serotec cat. #PIP014) at a final concentration of 1:20 for 48 hrs at 37°C/5%CO_2,_ as described [19]. Cell cultures for each subject included unstimulated wells (negative control) and phytohemagglutinin (PHA, 200ug/mL)-stimulated wells (positive control). Following incubation, supernatants were harvested and stored at −80°C until the electrochemiluminescence-based ELISA assays (Meso Scale Discovery, Rockville, MD) were performed. The coefficient of variation ranged from 10%-31% depending on the analyte [19].

### 2.3. Statistical analysis

The genotyping was completed using the Illumina Omni 1M array for subjects in the Rochester cohort (n=968), and using the Illumina 550 and 650 SNP arrays (for the European and African-American subjects, respectively) in the San Diego cohort (n=888) [17, 18, 22]. After thorough quality-control filtering (i.e., checks for duplicate samples; SNP call rates < 95%; sex reporting errors; departures from Hardy-Weinberg equilibrium p-value < 10^-6^; minor allele frequencies <0.005; indel/multi-allelic/monomorphic SNP removal), 964 Rochester and 807 San Diego subjects met the standards for imputation input. A set of >6 million SNPs was imputed using the University of Michigan Imputation Server [23] and the Haplotype Reference Consortium (for subjects with non-African ancestry) for each array. After thoroughly evaluating the quality of the genotype data (removal of duplicate SNP basepairs, replacing imputed SNPs with observed counterparts when available), we used the genetic data to define major ancestry groups. Because of constraints on PBMC samples and corresponding availability of cytokine/chemokine secretion data, we restricted out analysis to the European/Asian ancestry groups. Genetic ancestry was identified using the STRUTURE software with 1000 Genome reference panels and was performed on the combined 1,771 self-identified non-African American subjects. SNPs with r^2 imputation quality >=0.8 were used, with 585 1000 Genome samples included as anchors. Based on STRUCTURE metrics, 11 of these subjects were identified as having a high likelihood of African ancestry (probability >0.5) and were removed from further analysis. Of the remaining 1,760 subjects, cytokines/chemokines secretion were measured for 1,406 subjects who had samples available. Among these 1,406 subjects, 79 subjects were identified to have Asian ancestry. Relationship checking was performed using identical by descent (IBD) estimations from PLINK for all pairs of subjects. Full siblings and parent-offspring relationships were identified. To account for related pairs in the analyses, we estimated the variance of the association statistics by using the IBD information, as described in Schaid et al. [24]. Principal components were calculated to adjust for population stratification.

Our initial GWAS of mumps-specific cytokines/chemokines was focused on the n=1,406 (903 males and 503 females) subjects with European or Asian ancestry (i.e., Rochester cohort, n=748 subjects, San Diego cohort, n=658 subjects). The covariates that were significantly associated with the cytokine/chemokine trait were the following: age at last MMR vaccination; cohort (San Diego vs. Rochester); sex; age at blood draw; year of blood draw and the first 10 principal components (PCs) that were constructed from the genome wide SNP arrays. These associated covariates were used as independent variables in linear models with quantile-normalized cytokine/chemokine trait as dependent. The residuals from these models were then used as the dependent in each SNP association (measured and imputed). Genotypes were included in the models as the dose of the minor allele. To be assessed, SNPs needed both an imputation r-squared quality of at least 0.3 and minor allele frequency between 0.01 and 0.99 across all three arrays. In total, 7,338,443 SNPs were analyzed. Linear modeling was conducted using R software version 3.5.2.

## 3. Results

### 3.1. Study sample characteristics

The analyzed cohort consisted of subjects with European ancestry (EUR, 94.4%) or Asian ancestry (5.6%). The sex distribution was 903 males (64.2%) and 503 females (35.8%). The average age at enrollment was 19.5 years (Table 1). The mumps-specific background-subtracted cellular immune responses for the entire cohort are listed as medians and include the ranges in Table 2. Median values ranged from 2.95pg/ml (IQR, 0.79-6.78) for IL-10 to 5,756.28pg/ml (IQR, 2,265.37-10,766.02) for MCP-1 (Table 2).

**Table 2.** Cytokine/chemokine characteristics of the study subjects (n=1,406 subjects).

| **Cytokine/chemokine**<br>**(pg/ml)** | **Range** | **Median** | **Interquartile range,**<br>**IQR (25%, 75%)** |
| --- | --- | --- | --- |
| IFNα2 | -10.82 - 1931.85 | 24.13 | 1.81; 82.01 |
| IFNγ | -3050.7 - 2995.84 | 64.34 | 14.25; 176.94 |
| IL-10 | -11 - 116.78 | 2.95 | 0.79; 6.78 |
| IL-1β | -243.87 - 387.49 | 6.94 | 1.62; 20.39 |
| IL-2 | -54.3 - 326.04 | 10.97 | 4.07; 22.78 |
| IL-6 | -1114.69 - 2594.87 | 323.78 | 147.24; 679.76 |
| IP-10 | -4834.72 - 49177.41 | 434.62 | 81.7; 1407.68 |
| MCP-1 | -10065.15 - 36222.6 | 5756.28 | 2265.37; 10766.02 |
| MIP-1α | -5972.89 - 19906.55 | 528.41 | 179.25; 1375.39 |
| MIP-1β | -3112.78 - 6522.93 | 293.33 | 74.47; 694.57 |
| TNFα | -134.5 - 716.79 | 31.53 | 13.08; 65.2 |
| Results are reported as raw background-subtracted for cytokine/chemokine secretion values. The mumps-specific cytokine/chemokine immune responses for our study subjects reflect a Th1/pro-inflammatory response. |  |  |  |

### 3.2. Genome-wide analysis results with mumps cellular/inflammatory immune responses SIGLEC genomic region findings

Eleven cytokine and chemokines were selected for study based on prior literature indicating a role in mumps or paramyxovirus-specific immune responses [25-28]. Of these, four (IFNγ, IL-2, IL-1β, and TNFα) illustrated in Figure 1A-D demonstrated GWAS signals reaching genome-wide significance (p<5E-08). These panels illustrate the Manhattan plots of the –log10 (p-value) across the genome. MCP-1 secretion demonstrated a suggestive signal in the chromosome 13q14 region that did not quite meet the genome-wide significance threshold (p=5.45E-08) (Supplementary Figure 1). Supplementary Table 1 lists all SNPs with p-values 5.45E-08 to 7.01E-08 (including two coding rs1134071 and rs1926447; r2=0.73) associated with mumps-specific MCP-1 secretion.

**Figure 1A.**
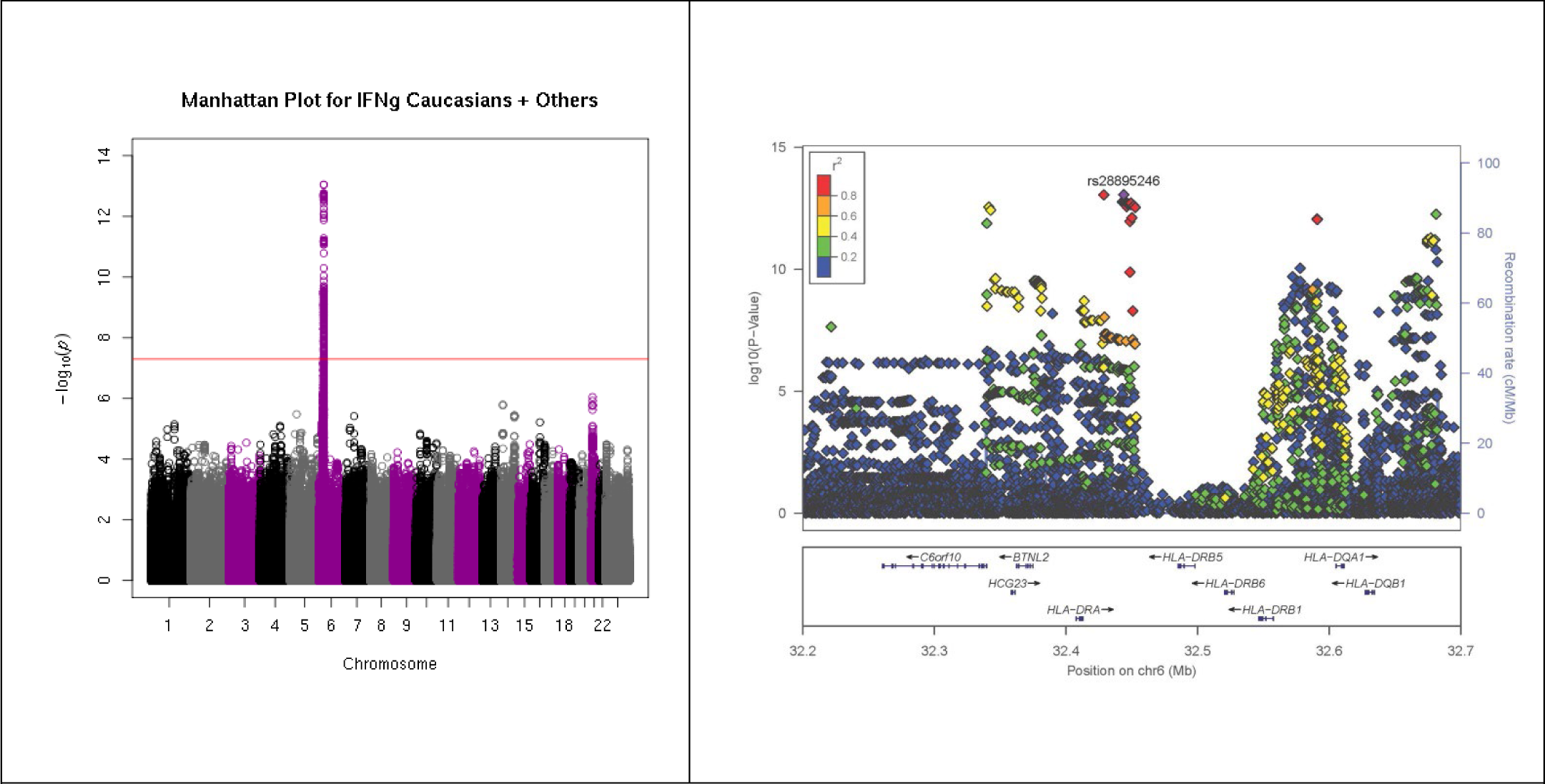
Genome-wide SNP associations with IFNγ secretion mumps vaccine recipients. A) Manhattan plot indicating SNPs associated with IFNγ response. The red line corresponds to p=5×10^-8^. B) Locus-zoom plot depicting region on chromosome 6p21 with the strongest association signal. SNP LD is shown in color. The name and location of each gene is shown at the bottom of the panel.

**Figure 1B.**
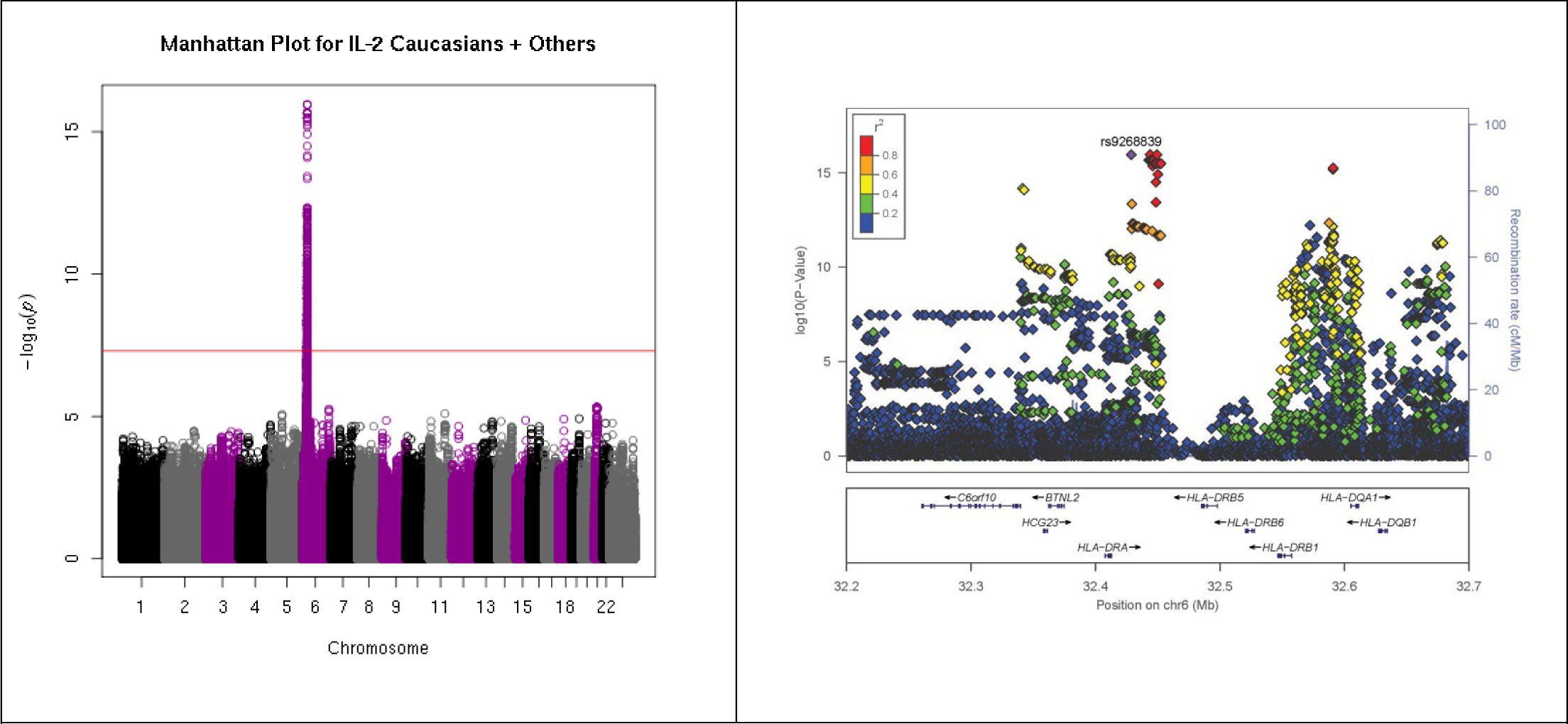
Genome-wide SNP associations with IL-2 secretion mumps vaccine recipients. A) Manhattan plot indicating SNPs associated with IL-2 response. The red line corresponds to p=5×10E^-8^. B) Locus-zoom plot depicting region on chromosome 6p21 with the strongest association signal. SNP LD is shown in color. The name and location of each gene is shown at the bottom of the panel.

**Figure 1C.**
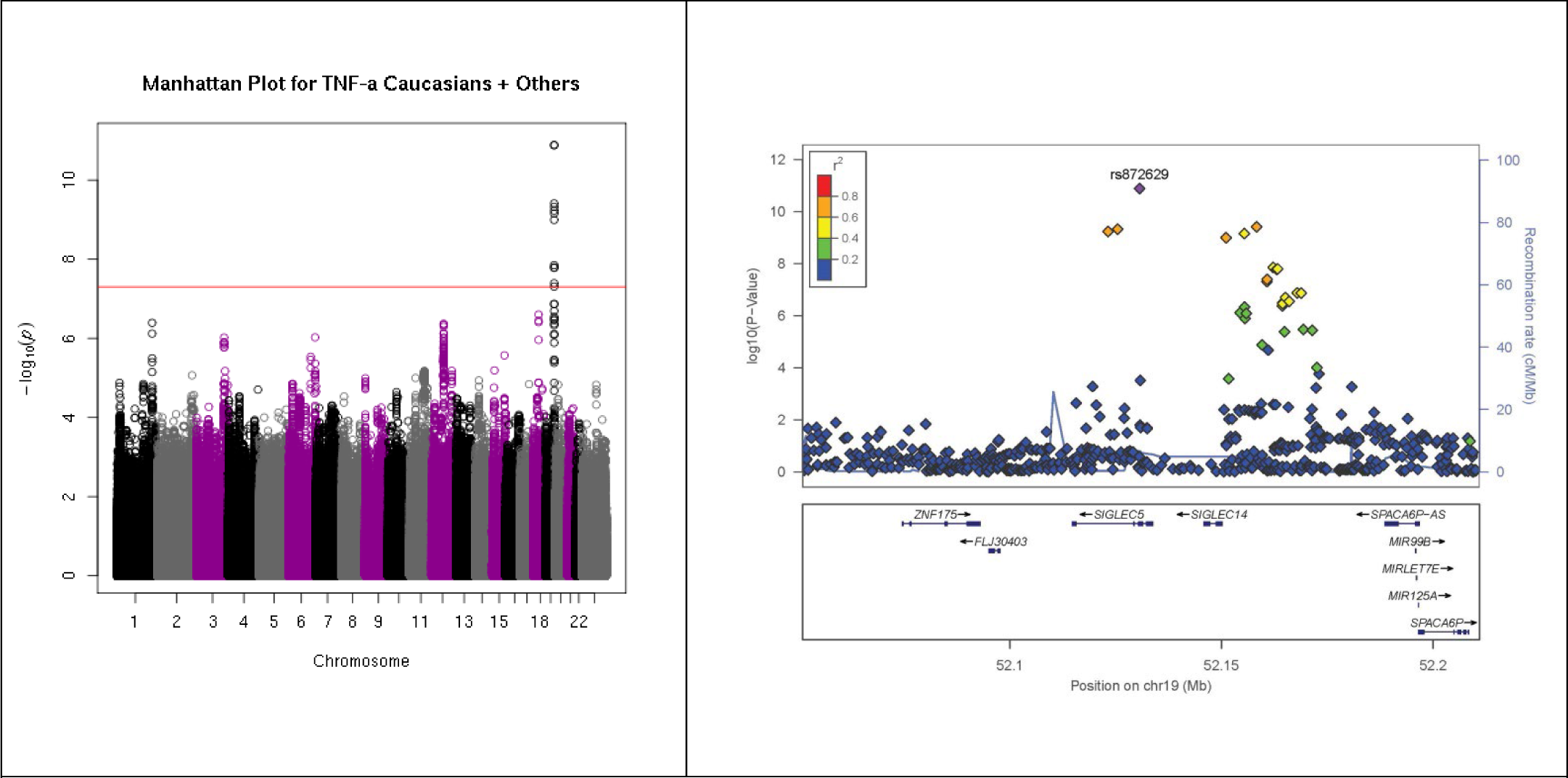
Genome-wide SNP associations with TNFα secretion mumps vaccine recipients. A) Manhattan plot indicating SNPs associated with TNFα response. The red line corresponds to p=5×10^-8^. B) Locus-zoom plot depicting region on chromosome 19q13 with the strongest association signal. SNP LD is shown in color. The name and location of each gene is shown at the bottom of the panel.

**Figure 1D.**
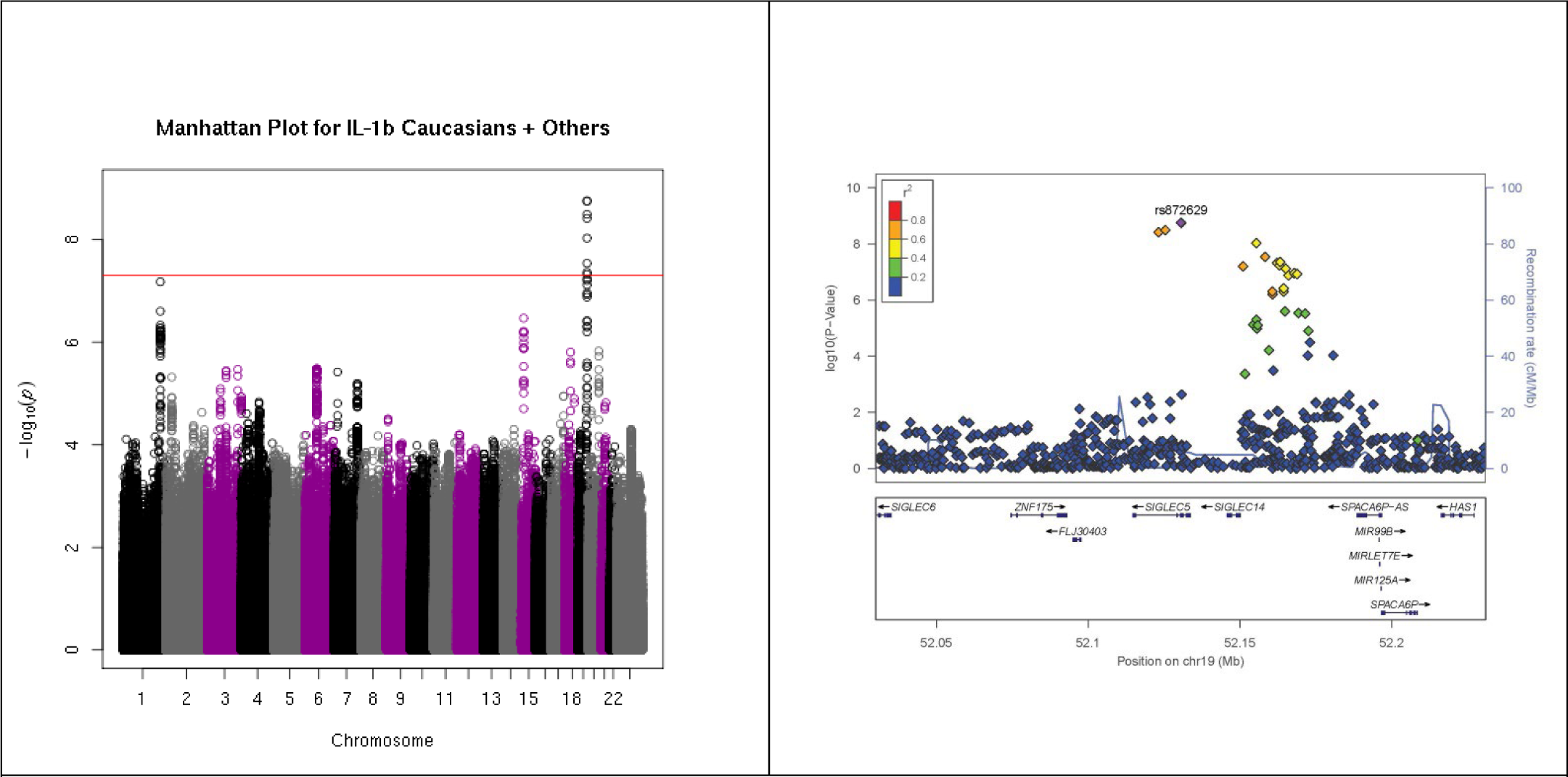
Genome-wide SNP associations with IL-1β secretion mumps vaccine recipients. A) Manhattan plot indicating SNPs associated with IL-1β response. The red line corresponds to p=5×10E^-8^. B) Locus-zoom plot depicting region on chromosome 19q13 with the strongest association signal. SNP LD is shown in color. The name and location of each gene is shown at the bottom of the panel

A genomic region (SIGLEC) located on chromosome 19q13 was associated with IL-1β and TNFα responses. The genetic association signal from the 19q13 region was linked to a block of 14 genetic variants in and around the sialic acid binding Ig like lectin (SIGLEC5/SIGLEC14) and ribosomal protein L7 pseudogene 51(RPL7P51) genes. The most significant SIGLEC5 SNPs, rs872629 and rs1106476 (in high linkage disequilibrium, LD, r^2^=0.97, p=1.77E-09), rs112456998 (p=3.21E-09) and rs73050880 (p=3.85E-09, r^2^=0.93) lie in an intron of the transcript and were associated with variations in mumps-specific IL-1β secretion (Table 4, Figure 2A). The minor alleles (i.e. less frequent) (A) of rs872629 and rs1106476 were significantly associated with an allele dose-related decrease in mumps-specific IL-1β secretion (p<1.77E-09, p<1.78E-09).

**Figure 2.**
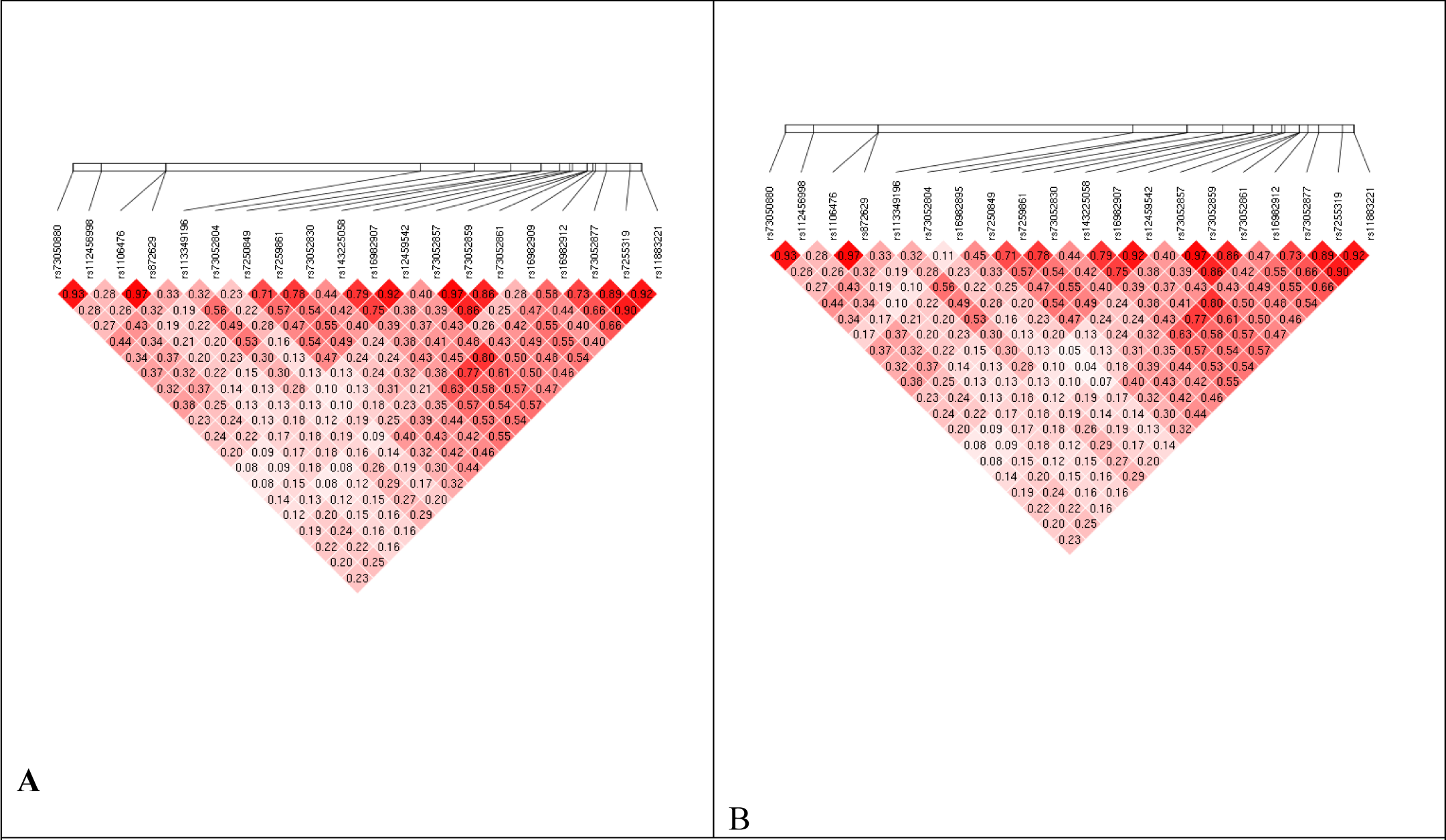
Haplotype block structure of the SIGLEC gene region in the study subjects. A). Linkage Disequilibrium (LD) patterns between SNPs associated with mumps-specific IL-1β secretion. B). Linkage Disequilibrium patterns between SNPs associated with mumps-specific TNFα secretion. The LD block structure was analyzed using Haploview software, version 3.32. The r2 color scheme is: white (r2=0), shades of red (0<r2<1), red (r2=1).

**Table 3.** HLA allelic associations with mumps-specific cytokine/chemokine immune responses.

| **Cytokine/**<br>**Chemokine** | **HLA**<br>**Locus** | **HLA Allele** | **Estimate** | **Std.**<br>**Error** | **Allele**<br>**frequency** | **P-value** |
| --- | --- | --- | --- | --- | --- | --- |
| IFNγ | DQA1 | DQA1*02:01 | 0.391899 | 0.063593 | 0.117823 | 9.45E-10 |
| IFNγ | DRB1 | DRB1*07:01 | 0.369561 | 0.0691 | 0.11877 | 1.05E-07 |
| IFNγ | DQB1 | DQB1*02:02 | 0.364409 | 0.068659 | 0.091263 | 1.30E-07 |
| IFNγ | DQA1 | DQA1*01:01 | 0.264266 | 0.064798 | 0.100036 | 4.80E-05 |
| IFNγ | DQB1 | DQB1*02:01 | -0.25401 | 0.063012 | 0.120687 | 5.86E-05 |
| IFNγ | C | C*06:02 | 0.315824 | 0.079431 | 0.073694 | 7.38E-05 |
| IFNγ | DQB1 | DQB1*05:01 | 0.243079 | 0.065131 | 0.109735 | 0.000198 |
| IFNγ | B | B*13:02 | 0.455027 | 0.131325 | 0.02027 | 0.000548 |
| IFNγ | DRB1 | DRB1*03:01 | -0.23984 | 0.069257 | 0.118461 | 0.000551 |
| IFNγ | DRB1 | DRB1*01:01 | 0.258569 | 0.07696 | 0.076448 | 0.000802 |
| IFNγ | DQA1 | DQA1*05:01 | -0.18841 | 0.060521 | 0.138107 | 0.00189 |
| IFNγ | B | B*08:01 | -0.21084 | 0.07008 | 0.114761 | 0.002675 |
| IFNγ | DQB1 | DQB1*03:03 | 0.269942 | 0.092815 | 0.044121 | 0.003693 |
| IFNγ | DRB1 | DRB1*08:03 | -0.7337 | 0.253809 | 0.004797 | 0.003906 |
| IFNγ | B | B*39:06 | -0.47196 | 0.177351 | 0.01069 | 0.007883 |
| IFNγ | C | C*07:01 | -0.16098 | 0.062024 | 0.152462 | 0.009552 |
| IL-2 | DQA1 | DQA1*02:01 | 0.433768 | 0.06698 | 0.117823 | 1.32E-10 |
| IL-2 | DRB1 | DRB1*07:01 | 0.445276 | 0.073022 | 0.11877 | 1.41E-09 |
| IL-2 | DQB1 | DQB1*02:02 | 0.419025 | 0.072388 | 0.091263 | 8.83E-09 |
| IL-2 | DQB1 | DQB1*02:01 | -0.27068 | 0.066434 | 0.120687 | 4.88E-05 |
| IL-2 | C | C*06:02 | 0.318497 | 0.083886 | 0.073694 | 0.000153 |
| IL-2 | DQA1 | DQA1*01:01 | 0.24203 | 0.068249 | 0.100036 | 0.000404 |
| IL-2 | DQB1 | DQB1*03:03 | 0.341808 | 0.097856 | 0.044121 | 0.000493 |
| IL-2 | B | B*13:02 | 0.447125 | 0.138499 | 0.02027 | 0.001276 |
| IL-2 | B | B*08:01 | -0.23381 | 0.073908 | 0.114761 | 0.001594 |
| IL-2 | DRB1 | DRB1*03:01 | -0.22686 | 0.073187 | 0.118461 | 0.001978 |
| IL-2 | DRB1 | DRB1*01:01 | 0.24928 | 0.081328 | 0.076448 | 0.00222 |
| IL-2 | DQA1 | DQA1*05:01 | -0.18333 | 0.063745 | 0.138107 | 0.00409 |
| IL-2 | DQA1 | DQA1*03:01 | 0.188875 | 0.067045 | 0.110545 | 0.004916 |
| IL-2 | DQB1 | DQB1*05:01 | 0.188891 | 0.068669 | 0.109735 | 0.006026 |
| IL-2 | DQA1 | DQA1*03:03 | 0.234328 | 0.085971 | 0.056499 | 0.006501 |
| IL-2 | C | C*04:01 | 0.187881 | 0.070332 | 0.118603 | 0.007647 |
| IP-10 | DRB1 | DRB1*15:05 | -0.99828 | 0.290994 | 0.00369 | 0.000621 |
| IP-10 | A | A*02:06 | -0.51637 | 0.181515 | 0.009637 | 0.004513 |
| MCP-1 | A | A*02:06 | -0.5519 | 0.173643 | 0.009637 | 0.001515 |
| MIP-1β | B | B*13:02 | 0.326345 | 0.121237 | 0.02027 | 0.007198 |
| IFNα-2α | DRB1 | DRB1*08:03 | -0.68224 | 0.218866 | 0.004797 | 0.001865 |
| IL-10 | C | C*06:02 | 0.223351 | 0.076274 | 0.073694 | 0.003466 |
| IL-6 | A | A*24:02 | 0.158327 | 0.058228 | 0.100136 | 0.006631 |

**Table 4.** Genome-wide significant associations of SNPs on chromosome 19q13 with mumps vaccine-specific IL-1β secretion (n=1,406 subjects).

| SNP ID<sup>a</sup> | bp | Minor allele | Major allele | Obs. 0<sup>b</sup> | Obs. 1<sup>b</sup> | Obs. 2<sup>b</sup> | MAF<sup>c</sup> | P-value | coeff | se | Gene | SNP location |
| --- | --- | --- | --- | --- | --- | --- | --- | --- | --- | --- | --- | --- |
| rs872629 | 52130638 | A | C | 1103 | 284 | 19 | 0.114509 | 1.77E-09 | -0.32401 | 0.011503 | SIGLEC5 | intron |
| rs1106476 | 52130637 | A | T | 1103 | 284 | 19 | 0.114509 | 1.78E-09 | -0.32386 | 0.011503 | SIGLEC5 | intron |
| rs112456998 | 52125447 | T | C | 1133 | 257 | 16 | 0.102774 | 3.21E-09 | -0.31048 | 0.011387 | SIGLEC5 | intron |
| rs73050880 | 52123194 | G | A | 1133 | 257 | 16 | 0.102774 | 3.85E-09 | -0.31138 | 0.011345 | SIGLEC5 | intron |
| rs73052804 | 52155401 | T | C | 1224 | 174 | 8 | 0.067568 | 9.36E-09 | -0.38688 | 0.009115 | SIGLEC14 | 5'upstream |
| rs7250849 | 52158316 | T | G | 1119 | 273 | 14 | 0.107041 | 2.9E-08 | -0.28726 | 0.011573 | SIGLEC14 | 5'upstream |
| rs12459542 | 52163278 | T | A | 974 | 396 | 36 | 0.16643 | 4.35E-08 | -0.23575 | 0.013932 | RPL7P51 | 3'downstream |
| rs143225058 | 52162236 | T | C | 973 | 397 | 36 | 0.166785 | 4.76E-08 | -0.23619 | 0.013891 | RPL7P51 | 3'downstream |
| rs16982907 | 52163035 | T | C | 973 | 397 | 36 | 0.166785 | 5.81E-08 | -0.23301 | 0.013952 | RPL7P51 | 3'downstream |
| rs113349196 | 52151087 | G | A | 1115 | 276 | 15 | 0.108819 | 6.29E-08 | -0.28961 | 0.011464 | SIGLEC14 | 5'upstream |
| rs16982912 | 52165127 | A | G | 951 | 413 | 42 | 0.176743 | 7.62E-08 | -0.22653 | 0.01426 | RPL7P51 | 3'downstream |
| rs7255319 | 52167906 | C | G | 956 | 408 | 42 | 0.174964 | 1.1E-07 | -0.22509 | 0.014198 | RPL7P51 | 3'downstream |
| rs11883221 | 52168832 | T | C | 956 | 408 | 42 | 0.174964 | 1.18E-07 | -0.225 | 0.014186 | RPL7P51 | 3'downstream |
| rs73052877 | 52165997 | T | C | 952 | 412 | 42 | 0.176387 | 1.32E-07 | -0.22295 | 0.014242 | RPL7P51 | 3'downstream |

Variant report tool was used to get annotations for SNPs of interest.
<sup>a</sup>SNP identification number
<sup>b</sup> Number of subjects with homozygous major allele genotype (0), heterozygous (1) and homozygous minor allele genotype (2)
<sup>c</sup> Minor allele frequency

Similarly, the most significant SIGLEC14 SNPs, rs73052804 (p=9.36E-09) and rs7250849 (p=2.9E-08, r2=0.23) were associated with variations in mumps-specific IL-1β secretion. Specifically, the minor alleles (T) of the 3’downstream rs73052804 and rs7250849 were significantly associated with an allele dose-related decrease in mumps-specific IL-1β secretion. We also identified statistically significant associations of rs73052804 (p=9.36E-09) and rs7250849 (p=2.9E-08, r^2^=0.23) in the SIGLEC14 genetic region with IL-1β cytokine response variations after mumps vaccination. All other statistically significant SNPs in this GWAS were in/near the RPL7P51 gene (Table 4).

The 19q13 region signal contained a block of SNPs in the SIGLEC5/SIGLEC14 genes associated with variations in TNFα secretion. The most significant SIGLEC5 SNPs, rs872629 and rs1106476 (in high LD, r^2^=0.97, p=1.3E-11) were associated with variations in TNFα secretion (Table 5, Figure 2B). The levels of IL-1β and TNFα were statistically associated with many common SNPs in strong LD. These SNPs included rs872629/rs1106476/rs112456998/rs73050880/rs73052804/rs7250849/rs12459542/rs143225058/rs169829 07/rs113349196/rs16982912/rs7255319/rs11883221 (IL-1β, r^2^>0.32; TNFa, r2>0.32). The Haploview output of variants within the SIGLEC gene associated with mumps-specific IL-1β and TNFα secretion are shown in Figure 2.

**Table 5.** Genome-wide significant associations of SNPs on chromosome 19q13 with mumps vaccine-specific TNFα secretion (n=1,406 subjects).

| SNP ID<sup>a</sup> | bp | Minor allele | Major allele | Obs.0<sub>b</sub> | Obs.1<sub>b</sub> | Obs.2<sub>b</sub> | MAF<sup>c</sup> | P-value | coeff | se | Gene | SNP location |
| --- | --- | --- | --- | --- | --- | --- | --- | --- | --- | --- | --- | --- |
| rs872629 | 52130638 | A | C | 1103 | 284 | 19 | 0.114509 | 1.3E-11 | -0.37884 | 0.011503 | SIGLEC5 | intron |
| rs1106476 | 52130637 | A | T | 1103 | 284 | 19 | 0.114509 | 1.32E-11 | -0.37867 | 0.011503 | SIGLEC5 | intron |
| rs7250849 | 52158316 | T | G | 1119 | 273 | 14 | 0.107041 | 3.89E-10 | -0.33692 | 0.011573 | SIGLEC14 | 5'upstream |
| rs112456998 | 52125447 | T | C | 1133 | 257 | 16 | 0.102774 | 4.76E-10 | -0.3395 | 0.011387 | SIGLEC5 | intron |
| rs73050880 | 52123194 | G | A | 1133 | 257 | 16 | 0.102774 | 5.9E-10 | -0.34034 | 0.011345 | SIGLEC5 | intron |
| rs73052804 | 52155401 | T | C | 1224 | 174 | 8 | 0.067568 | 6.98E-10 | -0.43196 | 0.009115 | SIGLEC14 | 5'upstream |
| rs113349196 | 52151087 | G | A | 1115 | 276 | 15 | 0.108819 | 1.02E-09 | -0.33983 | 0.011464 | SIGLEC14 | 5'upstream |
| rs143225058 | 52162236 | T | C | 973 | 397 | 36 | 0.166785 | 1.41E-08 | -0.25508 | 0.013891 | RPL7P51 | 3'downstream |
| rs12459542 | 52163278 | T | A | 974 | 396 | 36 | 0.16643 | 1.6E-08 | -0.25289 | 0.013932 | RPL7P51 | 3'downstream |
| rs16982907 | 52163035 | T | C | 973 | 397 | 36 | 0.166785 | 1.66E-08 | -0.25205 | 0.013952 | RPL7P51 | 3'downstream |
| rs73052830 | 52160765 | C | A | 1113 | 275 | 18 | 0.110597 | 4.06E-08 | -0.28756 | 0.011814 | RPL7P51 | 3'downstream |
| rs7259861 | 52160743 | A | C | 1093 | 293 | 20 | 0.118421 | 4.91E-08 | -0.278 | 0.012154 | RPL7P51 | 3'downstream |
| rs7255319 | 52167906 | C | G | 956 | 408 | 42 | 0.174964 | 1.35E-07 | -0.23236 | 0.014198 | RPL7P51 | 3'downstream |
| rs11883221 | 52168832 | T | C | 956 | 408 | 42 | 0.174964 | 1.36E-07 | -0.23275 | 0.014186 | RPL7P51 | 3'downstream |
| rs16982912 | 52165127 | A | G | 951 | 413 | 42 | 0.176743 | 2.03E-07 | -0.22764 | 0.01426 | RPL7P51 | 3'downstream |

Variant report tool was used to get annotations for SNPs of interest.
<sup>a</sup>SNP identification number.
<sup>b</sup> Number of subjects with homozygous major allele genotype (0), heterozygous (1) and homozygous minor allele genotype (2).
<sup>c</sup> Minor allele frequency.

### 3.3. HLA genomic region findings

We observed significant associations between HLA loci and cellular immune responses. The strongest associations were found in a genomic region on chromosome 6p21 (p<5E-08), associated with mumps-specific IFNγ and IL-2 secretion (Figure 1 A-B). The GWAS “top” SNP rs28895246 (C/T) in the HLA region was strongly correlated with mumps-specific IFNγ secretion (p-value=8.97E-14). The GWAS top SNP rs9268839 (A>C) in the HLA region was strongly correlated with IL-2 secretion (p-value=1.11E-16). This region was further analysed by using HLA genotype data (either measured or imputed by SNPs to 4-digit allele specificity) for HLA class I and II loci. HLA class IC, and HLA class II DQA1, DQB1 and DRB1 loci demonstrated suggestive associations (global p-value=1.00E-04) with mumps-specific IFNγ and IL-2 responses. In addition, the HLA class IB locus demonstrated a suggestive association with mumps-specific both IFNγ and IL-2 responses (p-value 0.001 to 0.0024) (Supplementary Table 2).

The HLA region was further analyzed using HLA genotyping data for major HLA loci. We found significant associations between mumps-specific IFNγ and IL-2 secretion and specific HLA class II (DQA1, DQB1 and DRB1) alleles (p-value=1.30E-07 to 1.32E-10); as summarized in Table 3. Several class II alleles (DQA1*02:01, DRB1*07:01 and DQB1*02:02, p<1.30E-07) were significantly associated with mumps-specific IFNγ production (Table 3). Regarding IL-2 secretion, HLA class II alleles (DQA1*02:01, DRB1*07:01 and DQB1*02:02, p<8.83E-09) were significantly associated with mumps-specific IL-2 production. HLA allelic p-values for other measured cytokines/chemokines (e.g., IP-10, MCP-1, MIP1β, IFNa2α, IL-10, and IL-6) ranged from 0.0066 to 0.00062 (Table 3).

## 4. Discussion

In this large cohort of mumps-vaccinated healthy individuals, we identified SNPs in two genetic regions that influence mumps vaccine-induced specific cytokine responses. One of our significant GWAS findings consisted of SNPs in high LD in the SIGLEC region on chromosome 19q13. Our study suggests that the SIGLEC5/SIGLEC14 gene SNPs play a role in the determination of mumps-specific IL-1β and TNFα secretion levels. Among the top significant SNPs in the SIGLEC region associated with mumps-specific IL-1β secretion are intronic SIGLEC5 rs872629, rs1106476, rs112456998, and rs73050880 (p=1.77E-09 to 3.85E-09) and 5’upstream SIGLEC14 rs73052804 and rs7250849 (p=9.36E-09 to 2.9E-08) (Table 4). The most significant SIGLEC5 SNPs, rs872629 and rs1106476 (in high LD, r2=0.97, p=1.3E-11) were also the top SNPs associated with variations in TNFα secretion (Table 5), with the majority of the other reported SNPs (Table 4, with the exception of rs73052877) also demonstrating an association with secreted TNFα. Thus, we report a signal/genetic variation in the SIGLEC5/SIGLEC14 genetic region, associated with, and likely impacting a common inflammatory pathway/signaling pathway after mumps vaccination.

SIGLECs are sialic acid-recognizing receptors of the immunoglobulin (Ig) superfamily and are important regulators of host immunity [29]. Sialic acid on the cell surface is known to be a common receptor for many paramyxoviruses as well as mumps virus. SIGLECs are known to help cell-cell interactions/signaling and control the functions of cells such as phagocytes, myeloid cells, macrophages, neutrophils, and monocytes, in innate and adaptive immunity via glycan recognition [29]. The inhibitory receptor Siglec-5 (encoded by *SIGLEC5*) has an immunoreceptor tyrosine-based inhibitory motif (ITIM), and is paired with the putative activating receptor Siglec-14 (encoded by *SIGLEC14*) bearing an immunoreceptor tyrosine-based activation motif (ITAM) motif, and these two proteins share >99% sequence homology in their first two ligand-binding Ig domains, but differ in function [29, 30]. Siglec-14 is demonstrated to interact with DAP12 to induce phosphoinositide-3-kinase (PI3K)/Akt pathway signaling and/or activation of other signaling cascades and promote proinflammatory response [31]. A deletion SIGLEC14 polymorphism has been described and is widespread in the human population (known as the SIGLEC14 “null” allele), resulting from ∼17 kb deletion and the fusion between SIGLEC5 and SIGLEC14 genes (presence of SIGLEC5-SIGLEC14 fusion gene) with lack of protein expression of functional Siglec-14 in homozygous “null”/minor allele individuals (the translated protein is identical in amino acid sequence to Siglec-5)[30, 32]. Importantly, the overexpression of Siglec-14 (the protein present in homozygous wild type allele and heterozygous individuals) in human monocytes has been demonstrated to significantly enhance TNFα and IL-8 secretion compared to Siglec-5 overexpression or control, demonstrating the functional consequences of this polymorphism [31, 32]. It is generally believed that the expression of polymorphic activating receptor SIGLEC14 with the inhibitory paired polymorphic receptor SIGLEC5 may balance immune/inflammatory responses to pathogens in sialic acids-SIGLEC dependent manner [31]. One of our top SIGLEC SNPs associated with TNFα/IL-1β secretion, rs1106476, has been previously identified as a proxy for the SIGLEC14 deletion/null allele polymorphism, with the minor AA genotype tagging the deletion/SIGLEC14 null phenotype (lack of Siglec-14 protein expression), that is also associated with increased expression of SIGLEC5/Siglec-5 [33]. Furthermore, the presence of the deletion/SIGLEC14 null phenotype has been associated with protection from exacerbations (commonly triggered by bacterial/viral infections) of chronic obstructive pulmonary disease/COPD, which is consistent with the diminished secretion of proinflammatory cytokines conferred by this genotype [34, 35]. Our GWAS results are interesting in light of studies showing that the trisaccharide-containing α2,3-linked sialic acid on the cell surface acts as a receptor for mumps virus and that α2,3-linked sialic acid is necessary for mumps-mediated cell-to-cell fusion and entry [36-39]. The functional regulation of Siglec family members is tightly controlled by the balance of *cis* (interactions with adjacent sialylated glycoconjugates on the same cell) and *trans* (with ligands on other cells or extracellular ligands) interactions, as well as by their unmasking from exposure to sialidases (such as cellular or pathogen-encoded sialidases) to confer different cell-to-cell interactions, cell activation/inhibition and cell signaling events [29, 40]. Our GWAS identified a significant association between the host inflammatory response to mumps (IL-1β and TNFα) and SIGLEC5/SIGLEC14 variants on chromosome 19q13. We can only speculate that the viral HN protein (on virions or infected cells) can regulate Siglec-5/Siglec-14 function and host inflammatory response by competition for ligand binding and/or by viral neuraminidase activity interfering with the masking/unmasking of Siglec-5/Siglec-14 on infected or neighboring cells [29, 40]. Of note, the glycosyltransferase *FUT2* gene (involved in host glycosylation and associated with mumps susceptibility in Tian et al., study) [41] is also located on chromosome 19q13. Therefore, we propose that a genomic region on chromosome 19q13 may affect mumps virus entry/susceptibility and innate/inflammatory response. In this regard, further research is needed.

The other significant SNPs associated with IL-1β secretion were located in/near the RPL7P51 gene. RPL7P51 codes for ribosomal protein L7 pseudogene 51, which plays a regulatory role in the mRNA translation. RPL7P51 has been shown to be an autoantigen and has yet to be linked to any immune function. We also identified 7 SNPs in the SIGLEC region and 5 SNPs in the RPL7P51 region that were significantly associated with mumps-specific TNFα response. Most SNP associations were observed in common with TNFα and IL-1β immune outcomes and demonstrated strong LD. Our results indicate that the genetic region encompassing SIGLEC (and RPL7P51) is associated with IL-1β and TNFα proinflammatory responses to mumps virus in vaccinated individuals and further functional mechanistic studies of these loci are warranted. Both IL-1β and TNFα are well recognized proinflammatory cytokines that are responsible for inducing a pyrogenic response to infection by working together with other cytokines [42].

Although our subjects demonstrated robust MCP-1 production in response to *in vitro* mumps virus stimulation, we did not identify any SNPs associated with this response at the genome-wide level of significance (p<5E-08). A cluster of SNPs located on chromosome 13q14 had p-values suggestive of an effect, including: intronic rs2182483 (p=5.45E-08) and coding rs1134071 (p=5.86E-08) in ZC3H13 (encoding for a zinc finger CCCH domain-containing protein 13) and intronic rs1926446 (p=6.36E-08) and coding rs1926447 (p=6.46E-08) in CPB2 (encoding a thrombin-activable fibrinolysis inhibitor, TAFI) (Supplementary Table 1). Published reports suggest that SNP associations not achieving a genome-wide level of significance may also be real and worth further study [43-45].

Genes/SNPs within the HLA region are major candidates for the regulation of adaptive immunity. Fine-mapping studies have linked variants in the HLA region to susceptibility to multiple infectious diseases [41]. In our earlier MMR vaccine studies, HLA alleles were associated with variations in immune responses to vaccines such as measles, mumps and rubella [46-48]. The Tian *et al.* GWAS study also reported the amino acid polymorphism HLA-A *Gln43* (rs114193679, p=2.51E-17) and the HLA-A*02:05 allele (p=1.73E-17) as a risk allele for susceptibility to mumps virus infection [41]. Our mumps GWAS found associations with cytokine production levels for several HLA alleles. Closer examination of the significant SNPs within the HLA region demonstrated that they are located within class II genes. In our subjects with European ancestry, the most significant HLA allele association with mumps-specific both IFNγ and IL-2 secretion was observed for DQA1*02:01, DRB1*07:01 and DQB1*02:02 alleles (Table 3). These results demonstrate that specific alleles at the DQA1, DQB1 and DRB1 loci show some level of association with cytokine immune responses, albeit at individual HLA locus association levels that do not achieve statistical significance (global p-value=1.00E-04).

There are no population-based studies identifying associations between mumps vaccine-induced immune responses and genome-wide SNPs, despite the immense public health implications. This study focused on identifying critical genetic variants associated with mumps vaccine-induced cellular immune responses by examining associations between SNPs polymorphisms and inter-individual variations in mumps-specific cytokine/chemokine immune responses. This provided the opportunity to examine not only immune genes likely to be involved in mumps vaccine response, but also the possibility of identifying the influence of new genes on mumps immunity. For example, GWAS studies have identified novel HLA and other genes impacting immune responses to vaccines including: measles [18], rubella [49, 50], hepatitis B [51], smallpox [17, 22], and others. As an example, the previously observed by Tian et al., genetic associations of rs516316 (p=9.63E-72), rs114193679 (p=2.23E-17), and rs3862630 (p=1.21E-08) in the FUT2, HLA-A Gln43, and ST3GAL4 genes, respectively, with susceptibility for mumps infection are also found within and outside of the HLA region [41]. Specific to mumps vaccine, Homan *et al.* attributed diminished protection to differential HLA presentation of T and B cell epitopes between the vaccine and wild type strains of mumps virus [52]. This diminished efficacy could theoretically be overcome by incorporating critical immunogenic epitopes into an improved vaccine strain virus.

Although our mumps vaccine GWAS study has many strengths including a well-defined and characterized cohort of two cohorts and a well-characterized viral vaccine, caveats are also present. We had a moderate sample size (<1,500) and limited numbers of diverse ancestries. Because susceptibility to many infectious diseases has been demonstrated to be influenced by genetic heterogeneity in different ancestral populations [53], sampling of other ancestries and ethnic groups and measuring mumps vaccine-induced immune response phenotypes could increase generalizability of our results. Our findings are based on measurements in subjects of European and Asian ancestry due to sample accessibility, thus future studies should include cohorts from more diverse ancestry. Some genetic variants may be false-positive associations with immune response outcomes and the association between specific HLA loci/alleles must be interpreted with caution and/or replicated in an independent cohort, followed by functional studies. However, we identified several genetic variants strongly associated with mumps vaccine-induced cytokine responses, including SIGLEC5/SIGLEC14.

In summary, our study is the first to identify common genetic variants associated with inter-individual variations in mumps-specific cytokine responses following live mumps vaccination. Although additional studies will be needed to validate these associations, our GWAS findings are an important step toward dissecting the host genetic influence on the immune response variation to live mumps vaccine. These studies could inform the development of novel mumps vaccine candidates and ultimately help combat waning immunity.

## Data Availability

All data produced in the present work are contained in the manuscript.

## Acknowledgments

We thank the Mayo Clinic Vaccine Research Group staff and the study participants. We are grateful to Dr. Steven A. Rubin and the Center for Biologics Evaluation and Research at the US Food and Drug Administration for their assistance in laboratory assays.

---

**Supplementary Table 1.** SNPs on chromosome 13q14 associated with mumps vaccine-specific MCP-1 secretion (n=1,406 subjects).

| SNP ID<sup>a</sup> | bp | Minor allele | Major allele | Obs.0<sup>b</sup> | Obs.1<sup>b</sup> | Obs.2<sup>b</sup> | MAF<sup>c</sup> | P-value | Coeff | se | Gene | SNP location |
| --- | --- | --- | --- | --- | --- | --- | --- | --- | --- | --- | --- | --- |
| rs2182483 | 46582600 | C | T | 658 | 620 | 128 | 0.311522 | 5.45E-08 | 0.200342 | 0.017111 | ZC3H13 | intron |
| rs1134071 | 46562935 | A | G | 671 | 609 | 126 | 0.306188 | 5.86E-08 | 0.196248 | 0.017206 | ZC3H13 | coding |
| rs1926446 | 46629995 | C | A | 693 | 591 | 122 | 0.296942 | 6.36E-08 | 0.195314 | 0.017153 | CPB2 | intron |
| rs1926447 | 46629944 | A | G | 693 | 591 | 122 | 0.296942 | 6.46E-08 | 0.195164 | 0.017155 | CPB2 | coding |
| rs992631 | 46591989 | C | T | 668 | 611 | 127 | 0.30761 | 6.82E-08 | 0.195571 | 0.017205 | ZC3H13 | intron |
| rs7998854 | 46592174 | C | A | 668 | 611 | 127 | 0.30761 | 6.85E-08 | 0.195545 | 0.017204 | ZC3H13 | intron |
| rs1319712 | 46591707 | T | C | 668 | 611 | 127 | 0.30761 | 6.87E-08 | 0.19552 | 0.017204 | ZC3H13 | intron |
| rs2404730 | 46617381 | G | A | 668 | 613 | 125 | 0.306899 | 6.9E-08 | 0.19624 | 0.017167 | CPB2 | 3'downstream |
| rs9526134 | 46618046 | G | T | 668 | 613 | 125 | 0.306899 | 6.97E-08 | 0.196168 | 0.017167 | CPB2 | 3'downstream |
| rs7331069 | 46564609 | A | T | 669 | 611 | 126 | 0.306899 | 6.99E-08 | 0.195141 | 0.017213 | ZC3H13 | intron |
| rs2404729 | 46617368 | G | A | 668 | 613 | 125 | 0.306899 | 7.01E-08 | 0.19614 | 0.017167 | CPB2 | 3'downstream |

Variant report tool was used to get annotations for SNPs of interest.
<sup>a</sup>SNP identification number.
<sup>b</sup> Number of subjects with homozygous major allele genotype (0), heterozygous (1) and homozygous minor allele genotype (2).
<sup>c</sup> Minor allele frequency.

**Supplementary Table 2.** HLA loci associations with mumps-specific IFNγ and IL-2 cytokine immune responses.

| **Cytokine** | **HLA Locus** | **Global P-value** |
| --- | --- | --- |
| IFNγ | HLA-C | 1.00E-04 |
| IFNγ | HLA-DQA1 | 1.00E-04 |
| IFNγ | HLA-DQB1 | 1.00E-04 |
| IFNγ | HLA-DRB1 | 1.00E-04 |
| IL-2 | HLA-C | 1.00E-04 |
| IL-2 | HLA-DQA1 | 1.00E-04 |
| IL-2 | HLA-DQB1 | 1.00E-04 |
| IL-2 | HLA-DRB1 | 1.00E-04 |
| IL-2 | HLA-B | 0.001 |
| IFNγ | HLA-B | 0.0024 |

**Supplementary Figure 1.**
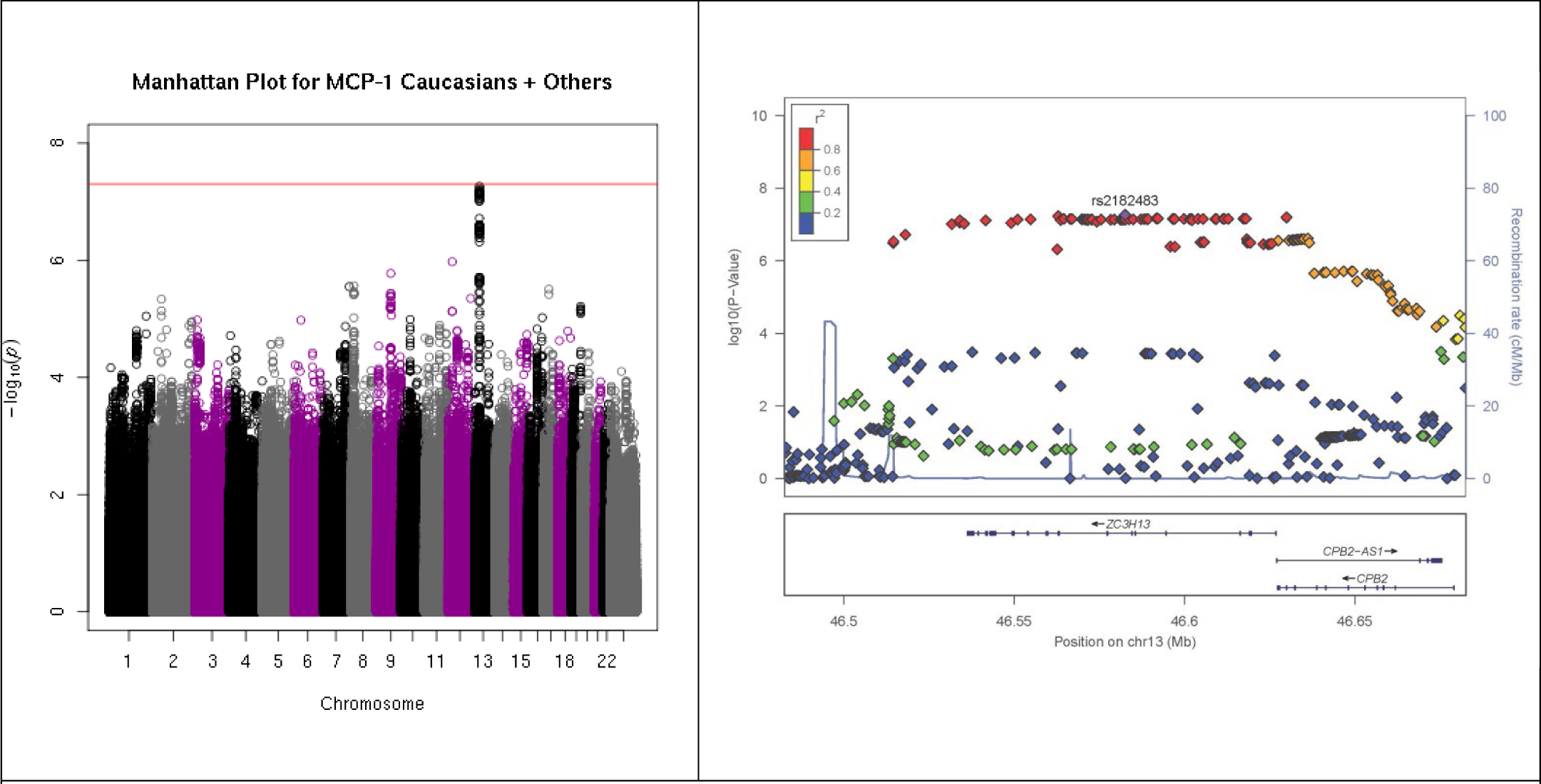
Genome-wide SNP associations with MCP-1 secretion mumps vaccine recipients. A) Manhattan plot indicating SNPs associated with MCP-1 response. The red line corresponds to p=5×10E-08. B) Locus-zoom plot depicting region on chromosome 13q14 with the strongest association signal. SNP LD is shown in color. The name and location of each gene is shown at the bottom of the panel.

## References

1. Dayan, G.H., et al., Recent resurgence of mumps in the United States. N.Engl.J Med, 2008. 358(15): p. 1580–1589.

2. Rubin, S.A., et al., Recent mumps outbreaks in vaccinated populations: no evidence of immune escape. Journal of Virology, 2012. 86(1): p. 615–20.

3. Grose, C., From King Nebuchadnezzar of Babylon to Mumps Genotyping and Vaccination 26 Centuries Later. Pediatrics, 2021. 148(6).

4. Brief report: update: mumps activity--United States, January 1-October 7, 2006. MMWR Morb.Mortal.Wkly.Rep., 2006. 55(42): p. 1152-1153.

5. Peltola, H., et al., Mumps outbreaks in Canada and the United States: time for new thinking on mumps vaccines. Clin Infect Dis, 2007. 45(4): p. 459–466.

6. Shepersky, L., et al., Mumps in Vaccinated Children and Adolescents: 2007-2019. Pediatrics, 2021. 148(6).

7. Dayan, G.H. and S. Rubin, Mumps outbreaks in vaccinated populations: are available mumps vaccines effective enough to prevent outbreaks? Clinical Infectious Diseases, 2008. 47(11): p. 1458–1467.

8. Lam, E., J.B. Rosen, and J.R. Zucker, Mumps: an Update on Outbreaks, Vaccine Efficacy, and Genomic Diversity. Clin Microbiol Rev, 2020. 33(2).

9. De Laval, F., et al., Lower long-term immunogenicity of mumps component after MMR vaccine. Pediatr.Infect.Dis J, 2010. 29(11): p. 1062–1063.

10. Marin, M., et al., Mumps vaccination coverage and vaccine effectiveness in a large outbreak among college students--Iowa, 2006. Vaccine, 2008. 26(29-30): p. 3601-3607.

11. Cohen, C., et al., Vaccine effectiveness estimates, 2004-2005 mumps outbreak, England. Emerg.Infect Dis., 2007. 13(1): p. 12-17.

12. Albertson, J.P., et al., Mumps Outbreak at a University and Recommendation for a Third Dose of Measles-Mumps-Rubella Vaccine - Illinois, 2015-2016. MMWR. Morbidity and mortality weekly report, 2016. 65(29): p. 731-4.

13. Briss, P.A., et al., Sustained transmission of mumps in a highly vaccinated population: Assessment of primary vaccine failure and waning vaccine-induced immunity. Journal of Infectious Diseases, 1994. 169: p. 77–82.

14. LeBaron, C.W., et al., Persistence of mumps antibodies after 2 doses of measles-mumps-rubella vaccine. Journal of Infectious Diseases, 2008. 199(4): p. 552–560.

15. Tan, P.L., et al., Twin studies of immunogenicity - determining the genetic contribution to vaccine failure. Vaccine, 2001. 19: p. 2434–2439.

16. Ovsyannikova, I.G., et al., Human leukocyte antigen and cytokine receptor gene polymorphisms associated with heterogeneous immune responses to mumps viral vaccine. Pediatrics, 2008. 121(5): p. e1091–e1099.

17. Ovsyannikova, I.G., et al., Genome-wide association study of antibody response to smallpox vaccine. Vaccine, 2012. 30(28): p. 4182–4189.

18. Haralambieva, I.H., et al., Genome-Wide Associations of CD46 and IFI44L Genetic Variants with Neutralizing Antibody Response to Measles Vaccine.. Human Genetics, 2017. 136(4): p. 421–435.

19. Riggenbach, M.M., et al., Mumps virus-specific immune response outcomes and sex-based differences in a cohort of healthy adolescents. Clin Immunol, 2022. 234: p. 108912.

20. Kennedy, R.B., et al., Genome-wide analysis of polymorphisms associated with cytokine responses in smallpox vaccine recipients. Human genetics, 2012. 131(9): p. 1403–21.

21. Ovsyannikova, I.G., et al., Impact of cytokine and cytokine receptor gene polymorphisms on cellular immunity after smallpox vaccination. Gene, 2012. 510: p. 59–65.

22. Kennedy, R.B., et al., Genome-wide genetic associations with IFNgamma response to smallpox vaccine. Human Genetics, 2012. 131(9): p. 1433–1451.

23. Das, S., et al., Next-generation genotype imputation service and methods. Nat Genet, 2016. 48(10): p. 1284–1287.

24. Schaid, D.J., et al., Multiple genetic variant association testing by collapsing and kernel methods with pedigree or population structured data. Genetic Epidemiology, 2013. 37(5): p. 409–18.

25. Young, K.R., et al., Immunologic characterization of a novel inactivated nasal mumps virus vaccine adjuvanted with Protollin. Vaccine, 2014. 32(2): p. 238–45.

26. Otani, N., et al., Development of a simplified and convenient assay for cell-mediated immunity to the mumps virus. Journal of Immunological Methods, 2014. 411: p. 50–4.

27. Wang, W., et al., IL-6 and IFNgamma are elevated in severe mumps cases: a study of 960 mumps patients in China. Journal of infection in developing countries, 2014. 8(2): p. 208–14.

28. Sulik, A., et al., Increased levels of cytokines in cerebrospinal fluid of children with aseptic meningitis caused by mumps virus and echovirus 30. Scandinavian Journal of Immunology, 2014. 79(1): p. 68–72.

29. Crocker, P.R., J.C. Paulson, and A. Varki, Siglecs and their roles in the immune system. Nat Rev Immunol, 2007. 7(4): p. 255–66.

30. Angata, T., et al., Discovery of Siglec-14, a novel sialic acid receptor undergoing concerted evolution with Siglec-5 in primates. Faseb j, 2006. 20(12): p. 1964–73.

31. Ali, S.R., et al., Siglec-5 and Siglec-14 are polymorphic paired receptors that modulate neutrophil and amnion signaling responses to group B Streptococcus. J Exp Med, 2014. 211(6): p. 1231–42.

32. Yamanaka, M., et al., Deletion polymorphism of SIGLEC14 and its functional implications. Glycobiology, 2009. 19(8): p. 841–6.

33. Shaw, B.C., et al., Analysis of Genetic Variants Associated with Levels of Immune Modulating Proteins for Impact on Alzheimer’s Disease Risk Reveal a Potential Role for SIGLEC14. Genes (Basel), 2021. 12(7).

34. Angata, T., Associations of genetic polymorphisms of Siglecs with human diseases. Glycobiology, 2014. 24(9): p. 785–93.

35. Angata, T., et al., Loss of Siglec-14 reduces the risk of chronic obstructive pulmonary disease exacerbation. Cell Mol Life Sci, 2013. 70(17): p. 3199–210.

36. Kubota, M., et al., Trisaccharide containing α2,3-linked sialic acid is a receptor for mumps virus. Proc Natl Acad Sci U S A, 2016. 113(41): p. 11579–11584.

37. Kubota, M., et al., Molecular Mechanism of the Flexible Glycan Receptor Recognition by Mumps Virus. J Virol, 2019. 93(15).

38. Latner, D.R. and C.J. Hickman, Remembering mumps. PLoS Pathogens, 2015. 11(5): p. e1004791.

39. Yoshida, N., et al., Mumps virus reinfection is not a rare event confirmed by reverse transcription loop-mediated isothermal amplification. J Med Virol., 2008. 80(3): p. 517–523.

40. Crocker, P.R. and A. Varki, Siglecs in the immune system. Immunology, 2001. 103(2): p. 137–45.

41. Tian, C., et al., Genome-wide association and HLA region fine-mapping studies identify susceptibility loci for multiple common infections. Nat Commun, 2017. 8(1): p. 599.

42. Dinarello, C.A., Infection, fever, and exogenous and endogenous pyrogens: some concepts have changed. J Endotoxin.Res., 2004. 10(4): p. 201–222.

43. Panagiotou, O.A. and J.P. Ioannidis, What should the genome-wide significance threshold be? Empirical replication of borderline genetic associations. International Journal of Epidemiology, 2012. 41(1): p. 273–86.

44. Chen, Z., et al., Revisiting the genome-wide significance threshold for common variant GWAS. G3 (Bethesda), 2021. 11(2).

45. Hammond, R.K., et al., Biological constraints on GWAS SNPs at suggestive significance thresholds reveal additional BMI loci. Elife, 2021. 10.

46. Ovsyannikova, I.G., et al., Consistency of HLA associations between two independent measles vaccine cohorts: a replication study. Vaccine, 2012. 30(12): p. 2146–2152.

47. Ovsyannikova, I.G., et al., Human leukocyte antigen and cytokine receptor gene polymorphisms associated with heterogeneous immune responses to mumps viral vaccine. Pediatrics, 2008. 121 p. e1091–e1099.

48. Ovsyannikova, I.G., et al., Replication of rubella vaccine population genetic studies: validation of HLA genotype and humoral response associations. Vaccine, 2009. 27(49): p. 6926–6931.

49. Lambert, N.D., et al., Polymorphisms in HLA-DPB1 are associated with differences in rubella-specific humoral immunity after vaccination. Journal of Infectious Diseases, 2015. 211(6): p. 898–905.

50. Voigt, E.A., et al., Polymorphisms in the Wilms Tumor Gene Are Associated With Interindividual Variations in Rubella Virus-Specific Cellular Immunity After Measles-Mumps-Rubella II Vaccination. The Journal of Infectious Diseases, 2018. 217(4): p. 560–566.

51. Png, E., et al., A genome-wide association study of hepatitis B vaccine response in an Indonesian population reveals multiple independent risk variants in the HLA region. Human Molecular Genetics, 2011. 20(19): p. 3893–3898.

52. Homan, E.J. and R.D. Bremel, Are cases of mumps in vaccinated patients attributable to mismatches in both vaccine T-cell and B-cell epitopes?: An immunoinformatic analysis. Human vaccines & immunotherapeutics, 2014. 10(2): p. 290–300.

53. Burgner, D., S.E. Jamieson, and J.M. Blackwell, Genetic susceptibility to infectious diseases: big is beautiful, but will bigger be even better? Lancet Infect Dis, 2006. 6(10): p. 653–663.

